# Haptic Portable Robotic Device for Automated Guidewire or Catheter Navigation in Endovascular Procedures

**DOI:** 10.64898/2026.02.03.26345465

**Authors:** Vahid Mohammadi, Jason MacTaggart, Majid Jadidi, Alexey Kamenskiy

## Abstract

**Purpose:** Endovascular therapy is preferred over open surgery due to its minimally invasive nature, faster recovery, and lower perioperative risk; however, fluoroscopy guided procedures are limited by radiation exposure, high equipment costs, and reliance on highly skilled operators. This study aims to develop and evaluate a lightweight, portable robotic system for autonomous guidewire navigation to improve safety, accessibility, and operator independence.

**Methods:** A compact 400 g robotic device was designed with millimeter scale positioning accuracy, servo current based real-time haptic feedback, precise axial rotation, automated retraction advance control, and compatibility with standard endovascular tools. Miniature linear actuated servomotors replicate skilled manual maneuvers using impedance control. Upon tip contact, the system advances the guidewire by 1 mm, measures current changes, classifies lesion stiffness (soft, medium, stiff), and adapts virtual mass-spring-damper gains to regulate push speed and applied force. Bench experiments were conducted using flexible tubing with inserts simulating 20-80% stenosis and two current thresholds (75 mA and 94 mA).

**Results:** Retraction frequency increased with stenosis severity, validating the autonomous control strategy. Lesion stiffness classification achieved F-scores of 0.83, 0.77, and 0.95 for soft, medium, and stiff conditions, respectively, demonstrating reliable discrimination and adaptive force modulation.

**Conclusions:** The proposed system enables autonomous and adaptive guidewire advancement with high classification accuracy using low-cost, current based sensing and impedance control. Its lightweight and portable design reduces dependence on continuous manual operation and specialized imaging infrastructure, supporting safer and faster interventions and potential deployment in prehospital or resource-limited settings. This prototype advances the development of more accessible and operator-independent endovascular therapy.

## 1. INTRODUCTION

The automation of endovascular device advancement holds significant promise for improving procedural precision, reducing intervention times, and minimizing complications [1]. By standardizing catheter navigation, these technologies aim to make vascular procedures safer, more efficient, and less dependent on operator variability. In 2022 alone, nearly 9.5 million peripheral vascular procedures were performed in the U.S. [2], along with approximately 600,000 percutaneous coronary interventions [3]. Collectively, this amounts to more than 10 million percutaneous transluminal interventions annually. While generally effective, these procedures can be technically challenging - especially in patients with tortuous anatomy, severe calcification, or complex occlusions – scenarios that demand advanced operator skill and increase the risk of complications. This challenge is especially pronounced in catheter-based interventions such as mechanical thrombectomy for acute ischemic stroke and cardiac ablation for arrhythmias. Thrombectomy has significantly improved outcomes for patients with large vessel occlusions, yet only 5% of eligible patients receive the intervention due to transport delays, limited availability of trained specialists, and restricted access in non-tertiary centers [4]. Similarly, the complexity of catheter ablation has increased, with complication rates rising from 0.8% in 2005 to 1.8% in 2020 [5]. Although still low, this upward trend underscores the need for more consistent, precise, and assistive navigation technologies.

Robotic and automated navigation systems have emerged as promising solutions to these challenges. By enhancing catheter control, reducing reliance on manual dexterity, and enabling semi-autonomous or remote operation, such systems can improve procedural safety and reproducibility – particularly in time-critical or resource-limited environments. In trauma care, for example, robotic platforms could facilitate resuscitative endovascular balloon occlusion of the aorta (REBOA) in ambulances, rural hospitals, or even space missions – settings where trained personnel may be unavailable [6]. REBOA is a life-saving procedure for non-compressible torso hemorrhage, but its success hinges on expert skill for safe and timely placement. Automation could help overcome this barrier, expanding access in emergencies and mass-casualty incidents. A related and equally critical application is the treatment of traumatic aortic injuries – one of the leading causes of prehospital mortality, with approximately 80% of patients dying before reaching definitive care [7]. While open surgery was once the standard, it carried high risks due to invasive access and prolonged recovery. Thoracic endovascular aortic repair (TEVAR) has since become the preferred approach, reducing mortality to ∼7%, compared to 20–25% with open repair, and significantly lowering the risk of stroke and spinal cord injury [7]. TEVAR illustrates how minimally invasive techniques have revolutionized trauma care and exemplifies the value of automated technologies in high-risk vascular interventions.

More broadly, vascular interventional surgery (VIS) has replaced open surgery in many clinical areas by offering minimally invasive approaches that reduce trauma, shorten recovery, and improve outcomes. Standard VIS involves inserting a guidewire and catheter – often through a small incision in the wrist or groin – and navigating them under fluoroscopic guidance. Some catheters, such as ER-REBOA, Landmark REBOA, or COBRA-OS, bypass the guidewire step altogether [8]. Once positioned, devices like balloons, stents, or drug-delivery systems are deployed to treat the underlying pathology [9]. Despite their advantages, these procedures remain heavily dependent on operator experience. In high-risk trauma scenarios, manual device manipulation can be time-consuming and error-prone – delaying care when every second counts [10].

Recent advancements in robotic VIS have aimed to improve procedural precision, reduce complications, and lower radiation exposure, with commercial systems like Sensei and Magellan demonstrating early success [11]. However, these platforms remain limited by several critical shortcomings that prevent their widespread or emergency use. First, most existing systems are heavily dependent on manual operation by trained specialists and are typically confined to hospital-based, fixed installations. This severely limits their applicability in emergency or prehospital settings, where rapid response and wide accessibility are essential. Second, despite some integration of robotic control, current platforms lack true autonomy. Guidewire navigation continues to rely on operator skill, especially in anatomically complex or trauma-affected vessels. While machine learning and reinforcement learning frameworks - such as Soft Actor-Critic (SAC) - have shown potential in enabling semi-autonomous guidewire navigation using real-time imaging and feedback [12], [13], [14], these systems often require specialized catheters or hospital-grade imaging infrastructure, making them impractical for mobile or austere environments. Third, a major limitation is the lack of tactile sensing and real-time force feedback, which makes it difficult to detect contact with vessel walls. This increases the risk of vessel perforation and hemorrhage during navigation [15], [16], [17]. Moreover, most robotic systems lack a dedicated wire feeding mechanism capable of autonomously inserting and advancing the guidewire while adapting to vessel geometry and force conditions. As a result, guidewire manipulation remains a manual, labor-intensive task, adding procedural complexity and increasing the cognitive load on clinicians [18], [19], [20].

Together, these limitations reveal a fundamental gap: the field lacks a portable, intelligent, and autonomous endovascular system that combines precise guidewire manipulation with tactile awareness and real-time adaptability - especially one deployable beyond traditional hospital settings [21], [22], [23]. To address this need, we developed a portable robotic endovascular feeder designed to autonomously advance a guidewire or catheter using real-time force feedback and intelligent navigation algorithms. Unlike existing systems, our device features an integrated wire feeding mechanism capable of adjusting movement dynamically based on vessel interaction. Its compact, lightweight form factor allows rapid deployment in diverse environments - from modern hospitals to ambulances, field hospitals, or disaster zones. Portability enables remote operation, further expanding its utility in scenarios lacking on-site expertise. By integrating innovations in robotic catheter control, tactile sensing, and deep reinforcement learning, this platform offers a comprehensive solution to the limitations of current systems. It has the potential to reduce dependence on expert operators, enhance safety through adaptive control, and enable minimally invasive vascular interventions even in time-sensitive or resource-limited settings [24], [25].

## 2. MATERIALS AND METHODS

Traditional VIS involves several key procedural steps. The clinician begins by making a small incision - typically in the femoral (groin) or radial (arm) artery - to gain access to the vascular system. A guidewire is first inserted through this access point and carefully navigated toward the target location. As illustrated in Figure 1(a-d), the interventionalist manipulates the proximal end (tail) of the guidewire, often using one hand to advance and the other to rotate it, enabling directional control through the tortuous vasculature. Navigation is guided by real-time fluoroscopic imaging and, importantly, by the clinician’s tactile perception of resistance through the guidewire. When resistance increases - often due to vessel tortuosity, plaque buildup, dissection, or entry into an unintended branch - the guidewire is typically withdrawn slightly, rotated, and re-advanced to find a more favorable path. Once the guidewire reaches the intended site, a catheter or therapeutic device (e.g., balloon, stent, or drug delivery system) is advanced over the wire, which serves as a rail or track to guide the intervention [26], [27]. In anatomically complex lesions or tortuous vessels, this process can be highly demanding, even for experienced operators working under optimal hospital conditions with fluoroscopic guidance. The challenge becomes even greater in austere environments or when procedures are performed by non-specialist personnel, such as paramedics or battlefield medics, who may lack formal endovascular training. To address these limitations, we developed a robotic device that emulates the fine motor skills of an experienced interventionalist. The system reproduces both the linear and rotational motions used in manual guidewire manipulation and integrates tactile feedback to simulate resistance, thereby enabling safe and precise advancement of guidewires or catheters through complex vascular pathways.

**Figure 1.**
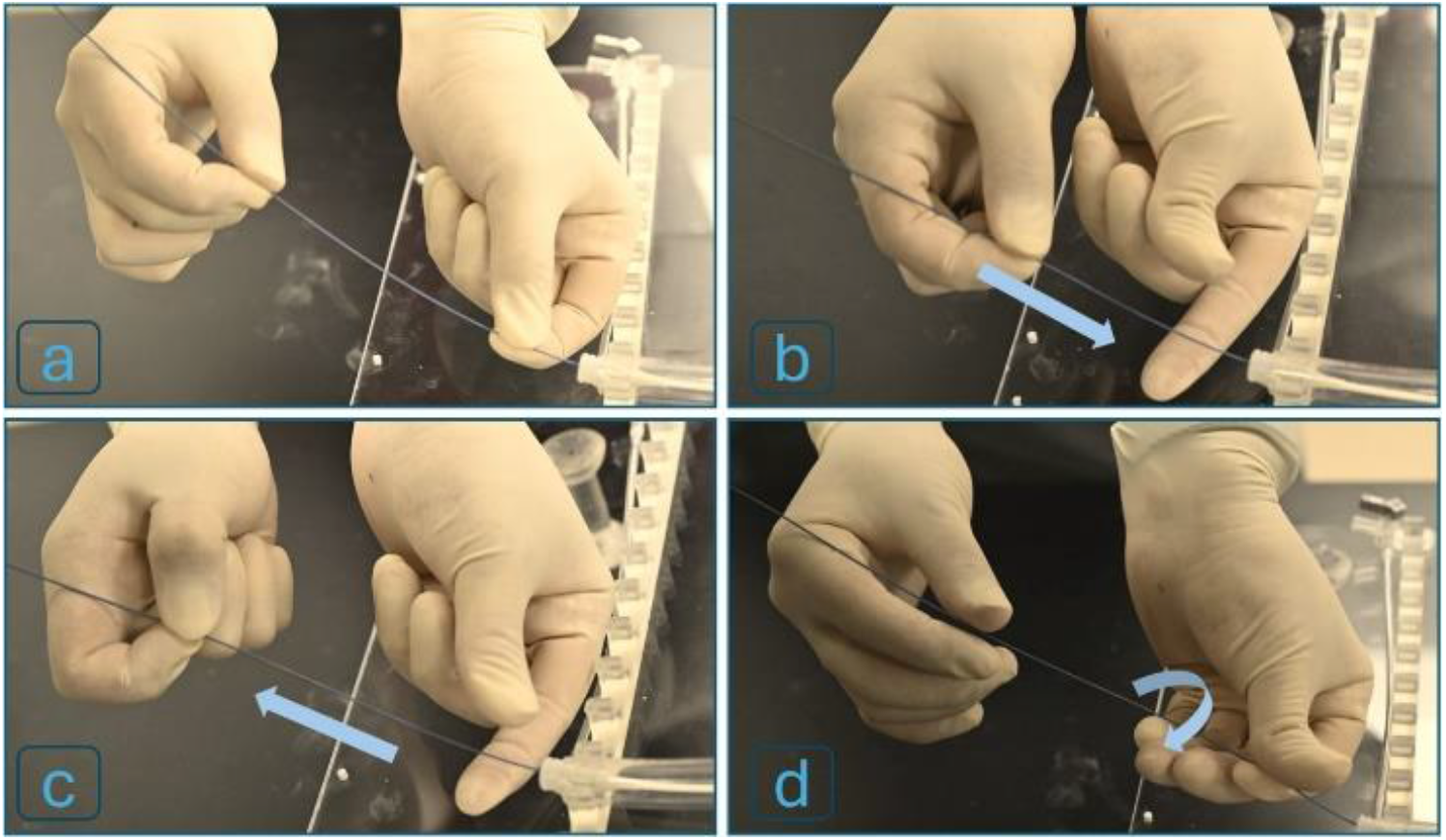
Key steps in traditional VIS, including (a) vascular access, (b) two-handed guidewire manipulation, (c) partial withdrawal and rotation of the guidewire upon sensing resistance, followed by re-advancement toward the target site, (d) rotation of the guidewire’s tail to facilitate advancement.

### 2.1. Mechanical Design

The robot’s mechanical structure was designed with an emphasis on compactness, portability, and versatility. A rear KST X06 servo motor (Servo 1) is responsible for gripping and advancing the wire, while a front fixed servo (Servo 2) controls rotational motion. A two-axis Dynamixel 2XC430 servo is used to drive the guidewire forward via a timing belt (first axis) and to rotate the entire mechanism when needed (second axis). An additional servo actuates one “finger” of the fixed gripper to twist the guidewire, mimicking the fine rotational motion typically performed by an interventionalist’s fingers (Figure 2(a-d)). At the device’s core is a lightweight, modular drive unit capable of advancing any standard catheter or guidewire. The system integrates all servo motors to replicate the essential motions used in manual guidewire manipulation (Figure 3a). Together, these components enable coordinated linear advancement and axial rotation, reproducing the two primary actions an operator uses to navigate a guidewire through the vasculature.

**Figure 2.**
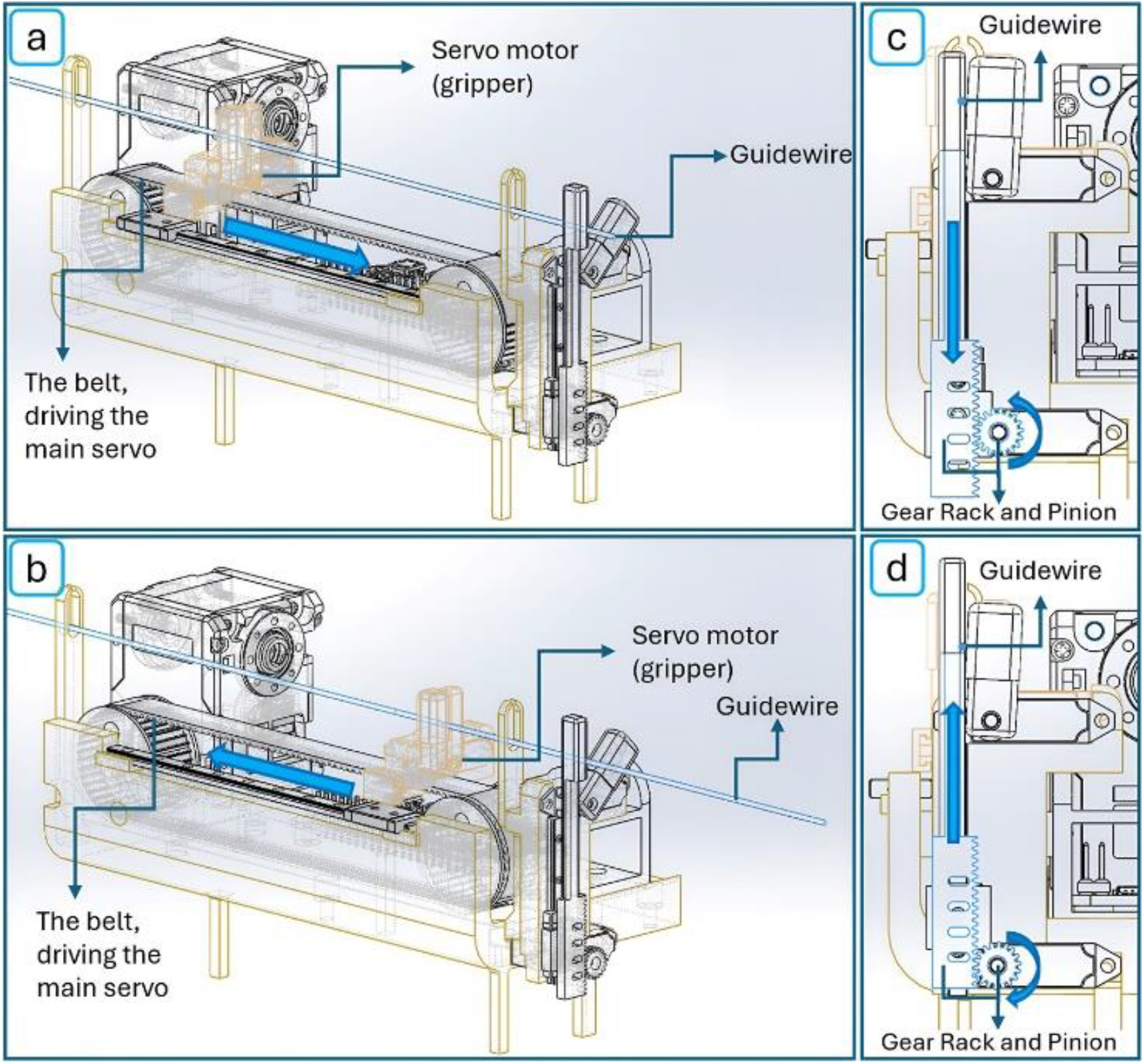
Robotic mechanism during guidewire manipulation: (a) servo motor advances the wire while the gripper is engaged, (b) the servo motor retracts after the gripper opens to prepare for the next advancement cycle, (c) the rack moves downward via a pinion rotated by a servo motor, mimicking human finger motion for twisting and rotating the wire, (d) the rack moves upward via the pinion.

**Figure 3.**
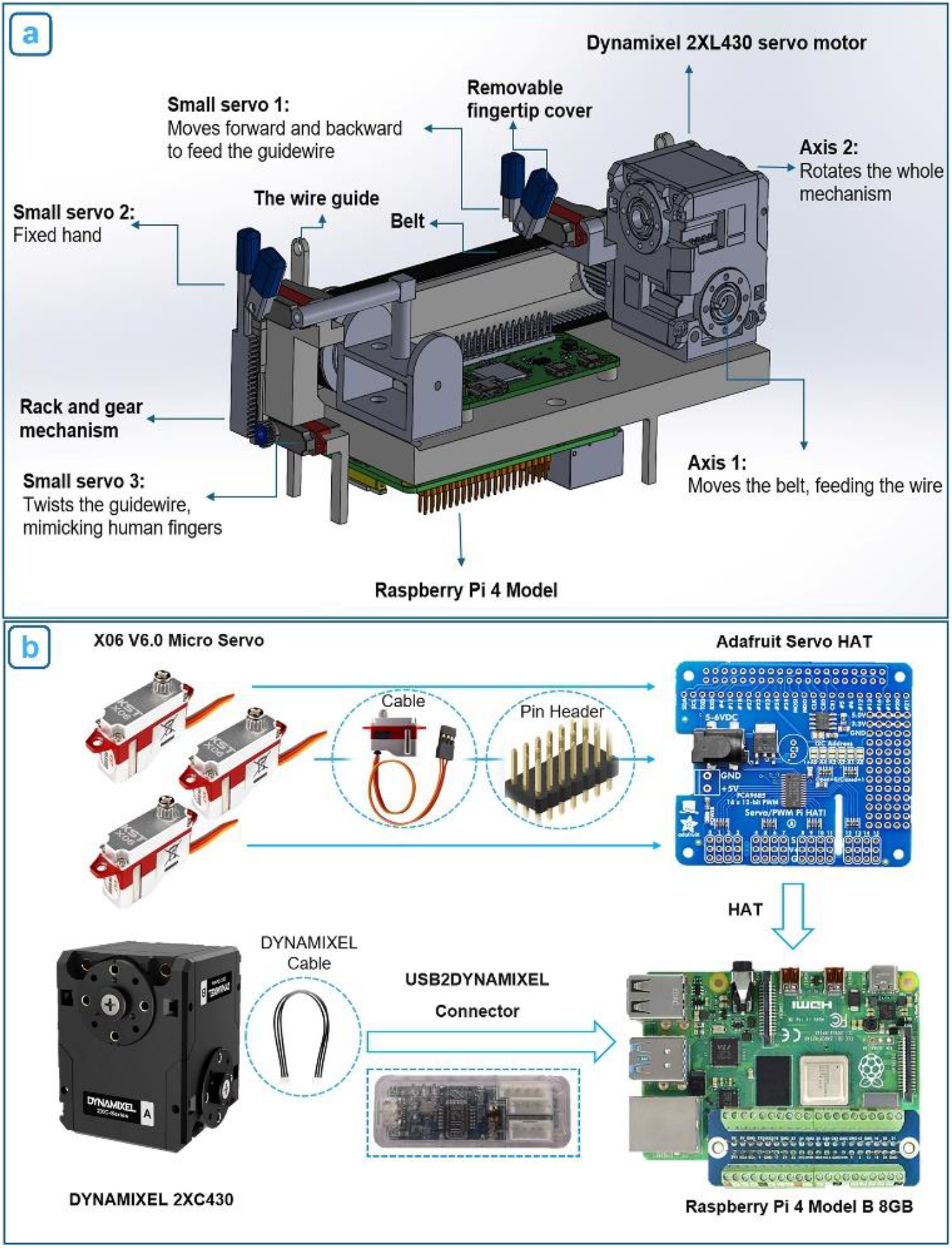
(a) Overview of the robot’s mechanical design, highlighting the main servo, three miniature servos, electronic control components, and key mechanical elements, including the guidewire advancement and rotation mechanisms, Mechatronic system schematic illustrates the connections between components. The three miniature servo motors are connected to the processor via a servo HAT, while the Dynamixel two-axis servo motor is interfaced with the processor using a communication converter

All components are mounted on a compact frame roughly the size of a mobile phone but with a thicker profile (160 mm × 75 mm × 90 mm), fabricated from SLA 3D-printed clear resin and durable ABS plastic. The structure includes two miniature linear guides that ensure high-precision motion control. The power transmission system for linear advancement of the guidewire or catheter consists of two pulleys and a belt, which are mounted on one of the linear guides to maintain stable forward and backward motion. For the twisting motion, a separate mechanism is employed: a rack-and-gear system, where the gear is driven by Servo 3, and the rack is mounted on a linear guide to stabilize the vertical movement of the robot’s movable “finger”. To prevent slippage during manipulation, flexible silicone fingertip covers are placed on each finger of both grippers. The main body also features two wire guides that help maintain horizontal alignment of the guidewire during advancement. Overall, this compact mechanical platform delivers the key motions required for vascular navigation - precise linear insertion and controlled axial rotation - in a highly miniaturized and portable form.

### 2.2. Mechatronic Architecture

The robot’s mechatronic architecture integrates precise actuation, sensor feedback, and control electronics to enable autonomous guidewire manipulation (Figure 3b). A two-axis Dynamixel 2XC430 servo motor with internal encoders provides both linear advancement and rotational control of the guidewire. This motor was selected for its compact size (36 mm × 46.5 mm × 36 mm), light weight (102 grams), and high torque output (stall torque: 1.8 N·m), enabling the robot to advance a guidewire through tortuous vasculature and perform fine rotational adjustments with high positional accuracy (resolution: 4096 pulses per revolution). The two-axis servo and the three miniature servo motors (KST X06) are connected to a Raspberry Pi 4 Model B (8GB), which serves as the central controller. The Raspberry Pi executes closed-loop control of motor speed and position using feedback from both motor encoders and a custom current-sensing circuit. To reduce electrical noise generated by the small servo motors, the system incorporates a custom-designed filter circuit that conditions the current signals before amplification and processing. These signals provide real-time information on resistance encountered during guidewire advancement, functioning as a tactile feedback mechanism. The entire system - including motors, control electronics, and the Raspberry Pi - is powered by a portable battery pack, enabling fully untethered operation. The robot is programmed using the Python language, which governs its autonomous behavior and real-time responsiveness to environmental conditions. This integrated mechatronic design allows the robot to autonomously feed and rotate a guidewire or catheter within the vasculature while monitoring resistance to avoid vessel injury. With built-in haptic feedback and programmable control logic, the system minimizes the need for user intervention until the guidewire reaches its target location.

### 2.3. Controller Design

#### 2.3.1. Finite-State Machine (FSM) Control

The FSM controller was designed to mimic how a skilled clinician responds to resistance during guidewire manipulation. It uses motor current as a proxy for contact force, allowing the robot to detect obstructions and respond in a human-like manner - by retracting, rotating, and re-advancing the guidewire to find a path forward [28], [29]. Force estimation is computed in real time using the formula:

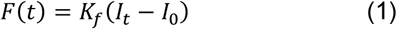

where *I*_*t*_ is the real-time current drawn by the feed motor, *I*_0_ is the no-load baseline current, and *K*_*f*_ is an experimentally determined calibration factor.

When the guidewire is advancing, the controller compares the estimated force *F*(*t*) to a predefined threshold *F*_*touch*_. If *F*(*t*)>*F*_*touch*_, feeding continues; if *F*(*t*)<*F*_*touch*_, the robot interprets this as a “touch event” and initiates a response routine. This allows the robot to mimic tactile sensing using only internal motor feedback – without external force sensors or complex modeling. As illustrated in Figure 4, the FSM control logic proceeds through the following steps:

- **Feeding phase:** Servo 1 advances the guidewire with smooth motion and low resistance.
- **Touch detection:** The system continuously monitors the current. When a peak exceeds the *F*_*touch*_ threshold, stenosis is detected, and a contact event is declared.
- **Whole-mechanism rotation (Option 1):** the guidewire is retracted 10 mm and rotated 90° via Servo 2 to reorient the tip.
- **Human-like twist (Option 2):** if resistance persists, the guidewire is retracted further, and Servo 5 actuates the movable “finger” via a rack-and-pinion mechanism to apply a twisting motion, similar to that performed by a human operator.

**Figure 4.**
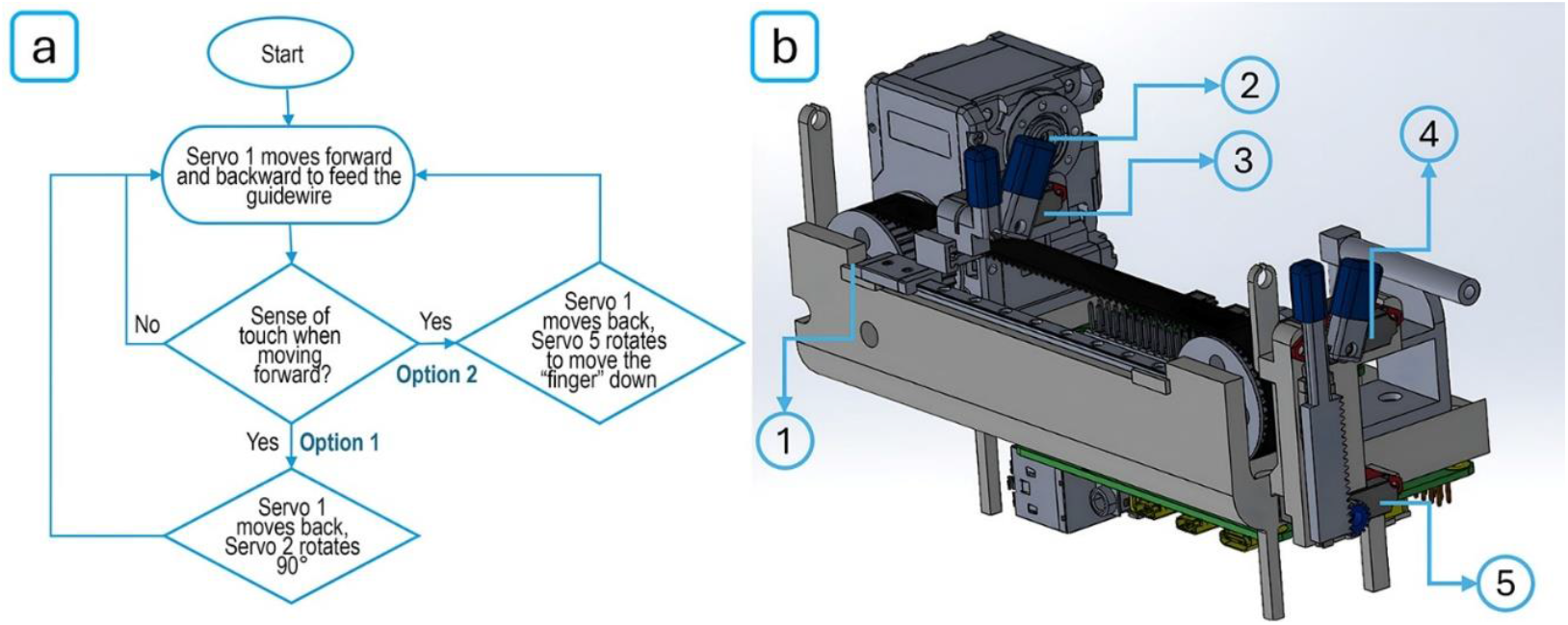
(a) Flowchart illustrating the Finite-State Machine (FSM) control logic used for autonomous guidewire navigation. The robot begins by advancing the guidewire (via Servo 1). If resistance is detected (interpreted as a sense of touch), it initiates one of two responses: Option 1-retract and rotate the guidewire 90° using Servo 2; or Option 2 - further retract and apply a human-like twist using Servo 5. The cycle repeats until advancement resumes; (b) Annotated rendering of the robotic system’s mechanical design showing key actuators: 1) Servo 1 (linear advancement), 2) Servo 2 (axial rotation), 3 & 4) guidewire feed mechanism with twisting gripper, 5) Servo 5 (twisting actuation via rack-and-pinion).

This rule-based framework enables adaptive, low-complexity behavior, minimizing the risk of excessive force that could lead to vessel dissection or perforation. The system continuously adjusts behavior based on internal force estimates, simulating the intuitive responses of an experienced interventionalist.

#### 2.3.2. Impedance Control and Force-Adaptive Navigation

While the FSM controller enables behavior-based navigation, it does not account for the variable stiffness of vascular stenoses. Lesions range from soft, lipid-rich plaques to rigid, calcified structures or organized thrombi. Navigating these lesions safely requires a controller that can adapt its motion profile in response to tissue compliance. To address this, we implemented an impedance control strategy, which allows the robot to behave like a mass–spring–damper system. This method enables the guidewire to move firmly through unobstructed vessels while becoming compliant when resistance is encountered [30], [31], [32]. The controller is governed by:

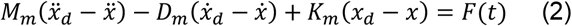

where *x*_*d*_ and x are the desired and actual positions of the guidewire tip; *M*_*m*_, *D*_*m*_, and *K*_*m*_ represent the virtual mass, damping, and stiffness parameters, respectively; and *F*(*t*) is the interaction force estimated from motor current. This formulation allows the robot to regulate both position and interaction force, smoothly adjusting its dynamics in response to the environment.

##### Stiffness Estimation

Prior to adjusting impedance, the robot must estimate the stiffness of the obstruction it has encountered. This is achieved through a brief two-step current measurement process:

1. Initial contact: At the moment of first contact (when a touch event is detected), record the motor current *I*_touch_.
2. Pressure application: Command a small forward displacement (we use 1 mm) and record the new motor current *I*_pressure_. If the guidewire cannot advance the full 1 mm due to a very stiff lesion, the actual displacement will be less.

The difference between these two current readings serves as a surrogate for obstruction stiffness. We define this current change as:

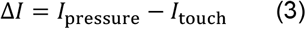

accompanied by the actual guidewire displacement during the 1 mm attempt:

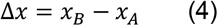

where *x*_*A*_ is the guidewire tip position at initial contact and *x*_*B*_ is the tip position after the attempted 1 mm advancement (Figure 5). Because the applied forward motion and time interval are fixed, a larger current increase Δ*I* (with smaller displacement Δ*x*) indicates a stiffer lesion, whereas a smaller Δ*I* (and larger Δ*x*) corresponds to a more compliant, soft lesion. In effect, we are measuring how much extra force is required to push the guidewire a short distance into the obstruction. This can be interpreted as an effective stiffness of the lesion:

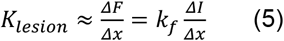

since Δ*F* ≈ *k*_*f*_Δ*I* from Equation (1). A nearly rigid blockage (high *K*_*lesion*_) will allow almost no forward movement (Δx≈0) for the given force, resulting in a sharp rise in current. In contrast, a soft plaque will deform or allow the guidewire to penetrate or deform, yielding a larger Δx with only a modest increase in current.

**Figure 5.**
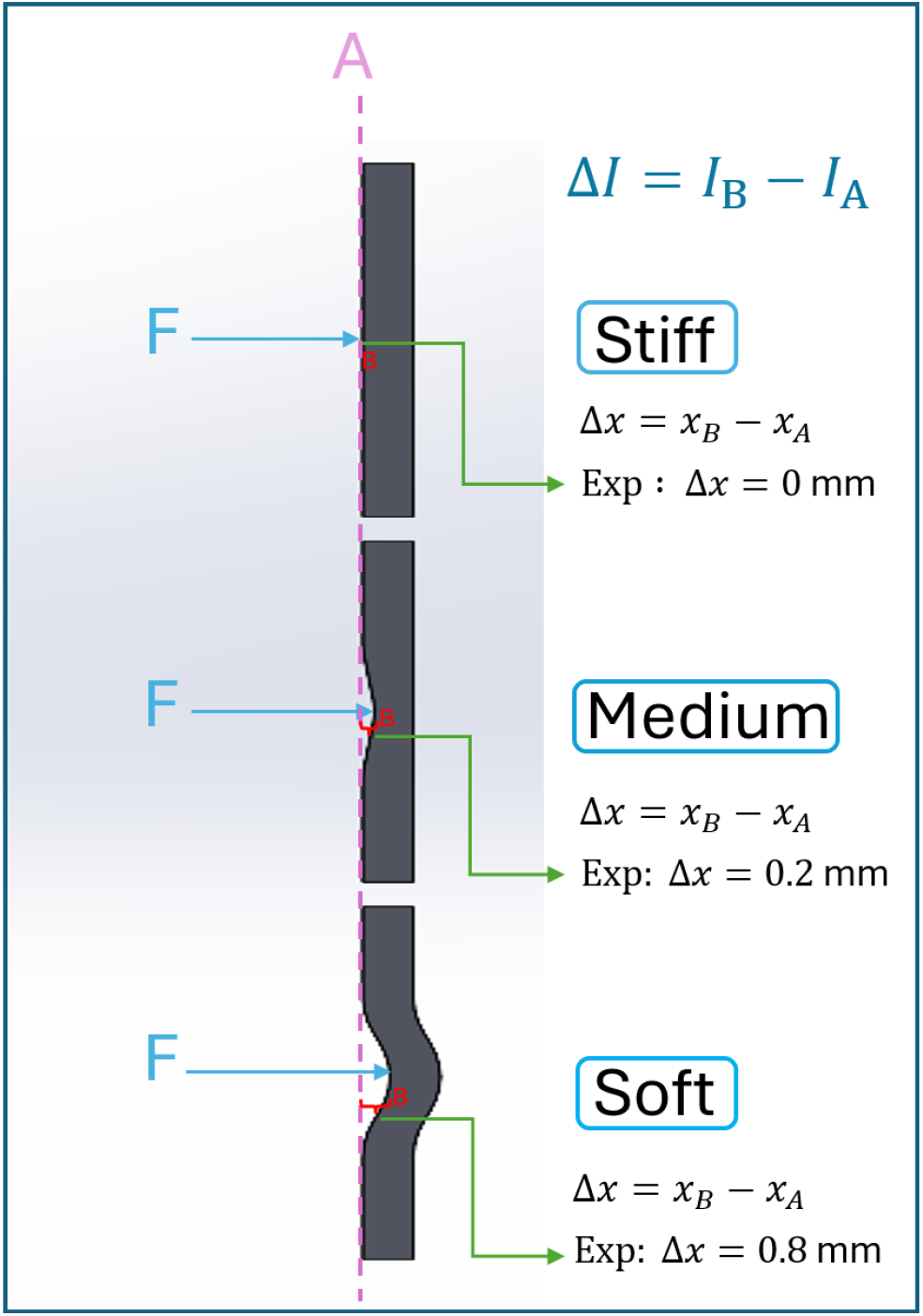
Classification of obstruction stiffness based on guidewire displacement and motor current feedback. When a constant force F is applied and the guidewire is advanced by 1 mm, the resulting displacement (Δx=x_B-x_A) and change in motor current (ΔI=I_B-I_A) depend on the compliance of the lesion. Stiff obstructions resist deformation, resulting in negligible displacement (Δx≈0) and a sharp increase in current. Medium obstructions allow limited deformation (Δx≈0.2 mm), producing a moderate current increase. Soft obstructions deform more readily (Δx≈0.8 mm), resulting in smaller current changes. This behavior enables the robot to estimate lesion stiffness in real time and adjust its impedance control parameters accordingly for safe and adaptive navigation.

Based on the measured *ΔI* (and inferred *K*_*lesion*_), the controller classifies the obstruction into one of three categories: Soft, Medium, or Stiff. For example, we can define two threshold values 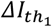 and 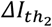 (determined experimentally) to delineate the categories:

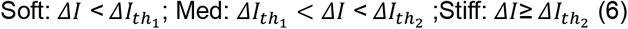

Bench tests showed that lesions could be reliably classified by ΔI: soft (0–5 mA), medium (5– 10 mA), and stiff (10–15 mA). This allowed real-time tuning of impedance parameters based on obstruction stiffness (So 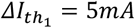 and 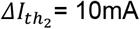).

##### Adaptive Impedance Adjustment

Once stiffness is classified, we adjust the virtual dynamics-*K*_*m*_, *D*_*m*_ and *M*_*m*_-based on the desired robot behavior. For soft lesions, the robot should move gently, so we reduce stiffness and increase damping. For stiff lesions, we aim for firmer and steadier motion, increasing stiffness and reducing compliance. This tuning allows the robot to adapt its response to different tissue types, mimicking human-like tactile behavior.

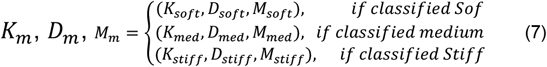

where each set of gains (*K*_*soft*_, *D*_*soft*_, *M*_*soft*_), etc., was predefined based on trial experiments to ensure stable yet responsive behavior. By adjusting these impedance parameters in real time according to feedback, the robot can advance the guidewire safely and adaptively. The mass-spring-damper behavior is tuned to avoid abrupt force spikes that might cause tissue damage: a higher damping *D*_*m*_ helps reduce overshoot and oscillations when resistance is met, and an appropriate virtual mass *M*_*m*_ provides stability during free-space navigation. This impedance control approach allows the system to traverse complex vasculature with human-like responsiveness, smoothly transitioning between rigid and compliant behavior in the presence of varying lesion stiffness.

### 2.4. Experimental Evaluation

To ensure measurement consistency and minimize biological variability, all experiments were conducted using an artificial artery model rather than excised vascular tissue. The artery was represented by a flexible silicone tube with an internal diameter of 15 mm, selected to approximate the geometry and compliance of the human aorta and iliac arteries. A modular experimental setup (Figure 6 (a-c)) was developed to allow repeatable positioning and controlled bending of the artificial vessel. The setup includes four identical mounting racks, each designed with a click-in slot mechanism that securely holds the tube in place. By repositioning the tube across different slots, various bending angles can be configured to simulate vascular curvatures seen in clinical scenarios. To further enhance experimental flexibility, a 5 × 5 positioning grid (25 total positions) was integrated into the test platform. This station rig allows the robotic system to be mounted at multiple locations around the simulated vessel, facilitating systematic evaluation of guidewire performance under different approach angles and spatial constraints.

**Figure 6.**
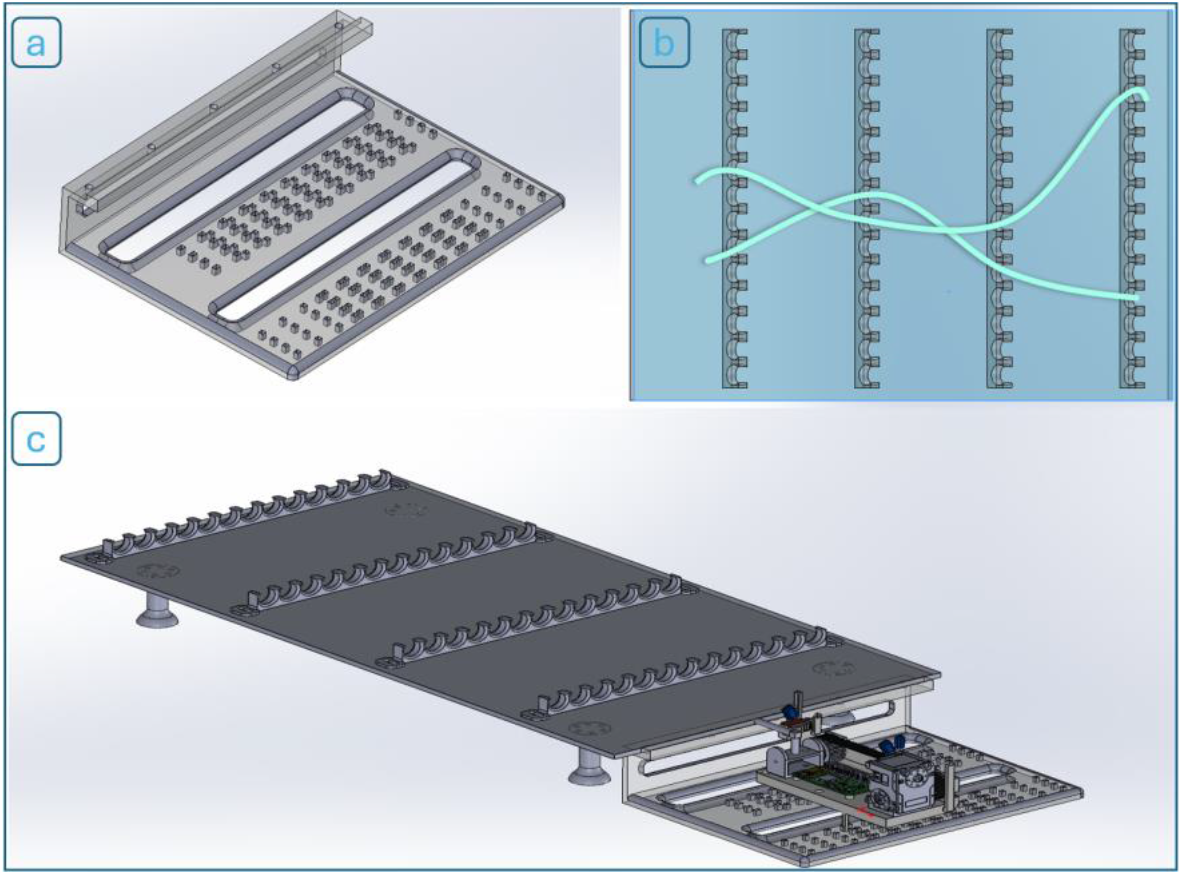
(a) Modular rack system and (b) two example bending paths in a 25-position station rig layout; (c) completely assembled testbed with the robot installed.

To simulate varying degrees of vascular stenosis, a series of custom-designed obstruction connectors was developed (Figure 7Figure 7). These 3D-printed inserts were manufactured to reduce the internal cross-sectional area of the flexible tube by 20%, 40%, 60%, and 80%, thereby mimicking mild to severe arterial narrowing. Each connector was made for easy insertion between two cut segments of the tube, allowing quick modification of the test environment. This modular design enables controlled and reproducible testing across different stenosis severities, replicating real-world anatomical challenges faced during endovascular procedures.

**Figure 7.**
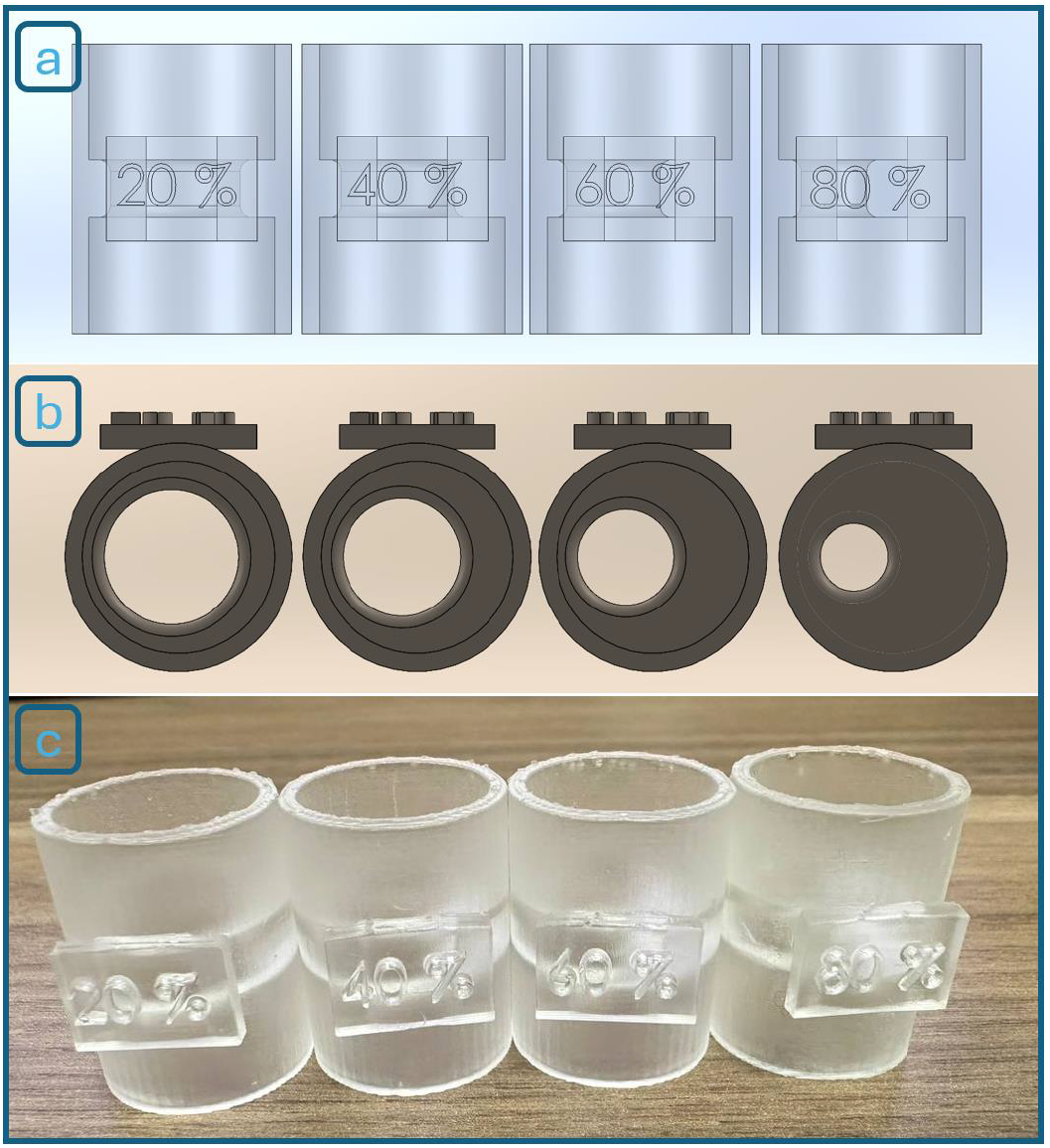
(a) Design of the obstruction connectors with (b) 20%, 40%, 60%, and 80% stenosis (left to right). (c) Physical 3D-printed components used in the experiments.

### 3 RESULTS

### 3.1 Volume of the flow lumen

Following the prototyping of all robotic components using FDM and SLA 3D printing, the robotic system (Figure 8a) and the complete experimental platform (Figure 8b) were assembled. A total of 80 experimental trials were performed, encompassing multiple stenosis levels and varying stenosis-detection sensitivity thresholds.

**Figure 8.**
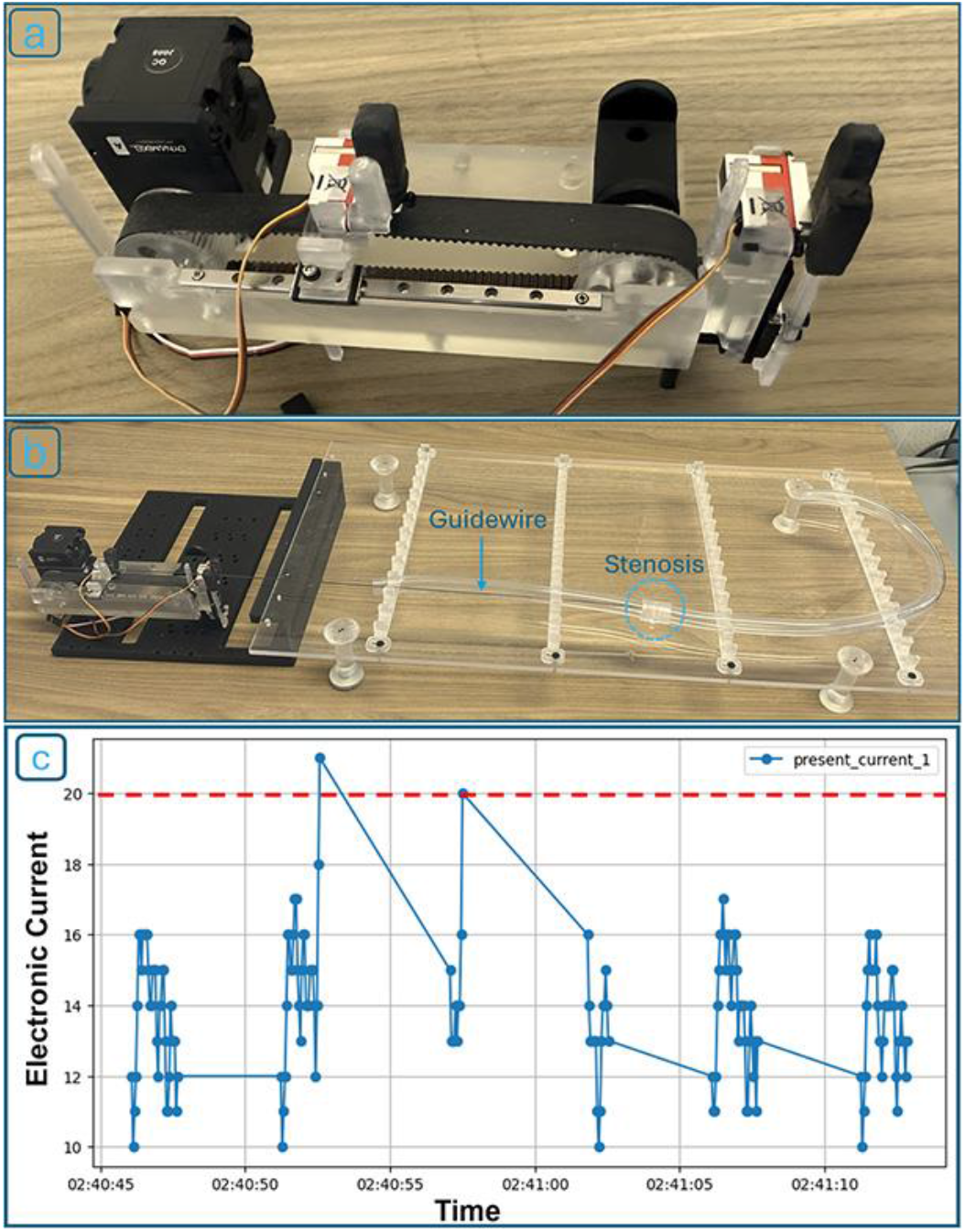
(a) Fully assembled robotic device; (b) robot positioned within the experimental setup, feeding the guidewire into the artificial artery; (c) Representative current profile from Servo Motor 1. Two distinct retraction events indicate obstruction detection and corrective guidewire maneuvers. The current detection threshold is depicted in red.

To begin each experiment, the robot was mounted at a selected position on the testing grid, and the flexible tube was bent to simulate vascular curvature. Once positioned, the robot was operated using a custom Python-based control program that continuously monitored the current drawn by the first axis of the Dynamixel servo motor. This motor current was used as a proxy for detecting obstructions, simulating the tactile feedback a human operator would feel. As the guidewire advanced, an increase in current indicated contact with either the arterial wall or a blockage. When a preset threshold was exceeded, the robot triggered a retraction and redirection routine, mimicking a clinician’s maneuver to bypass an obstruction. Figure 8c shows a representative current vs. time graph for one trial, where two such retractions occurred in response to obstruction events.

### 3.2 Effect of Stenosis Severity on Robot Performance)

The main performance metric in these trials was the number of retractions required for the robot to navigate through different stenosis levels. We tested four obstruction severities (20%, 40%, 60%, and 80% cross-sectional reduction), with 10 trials conducted per level, totaling 40 experiments. For each trial, we recorded the number of retractions until successful passage through the simulated lesion. We repeated the full set of 40 trials under two obstruction-detection sensitivity settings, defined by different motor current thresholds: 94 mA (baseline) and 75 mA (higher sensitivity). In the next phase, we repeated all these experiments using obstruction parts printed with flexible material (Formlabs Flexible Resin 80A, thickness 2.5mm). This brought the total number of experiments to 160, covering three variables: stenosis severity, sensitivity level, and obstruction material stiffness.

Figure 9a shows bar plots of average retractions per stenosis level (rigid) at 94 mA, while Figure 9b provides a violin plot that reveals the full distribution, density, and variability of retraction counts across trials. As stenosis severity increased, the number of retractions rose accordingly. At 20% stenosis, the robot passed with no or one retraction in 7 out of 10 trials. At 80%, all trials required at least one retraction, and 3 trials required four or more. The violin plot confirmed this upward trend, with both median and variability increasing with stenosis level.

**Figure 9.**
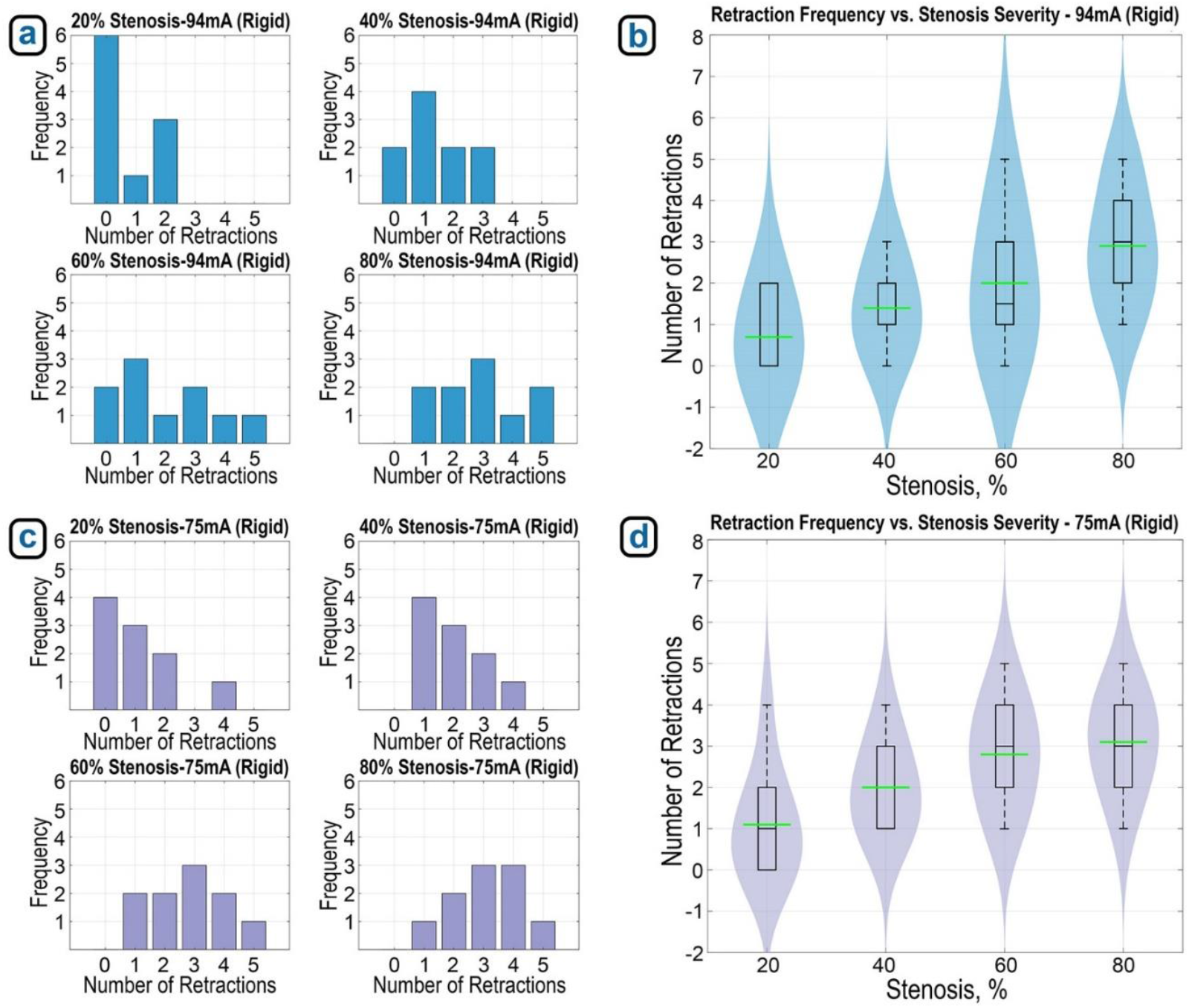
(a) Average number of retractions at each stenosis severity level (rigid, 10 trials each) with 94 mA detection threshold, and (b) violin plot showing distribution and density of retractions across stenosis severity under this threshold. Average retractions per stenosis severity (rigid, 10 trials each) at 75 mA threshold, and (d) violin plot showing distribution of retractions across stenosis severity with increased sensitivity.

Repeating the experiments at a more sensitive threshold (75 mA, Figure 9c) resulted in higher retraction counts across all stenosis levels. At this lower threshold, the robot triggered retractions even in less severe stenosis. Across all obstruction levels, the average number of retractions exceeded two. In 80% stenosis trials, the robot required up to five retractions in some cases (Figure 9d), confirming that increased obstruction detection sensitivity comes at the price of increased retraction frequency.

We also obtained similar data for the flexible material at different stenosis severity levels, as shown in Figure 10. Figure 10a demonstrates that the number of retractions increased with greater stenosis severity. At 20% stenosis, the robot passed with zero or one retraction in 8 out of 10 trials. At 80%, 9 out of 10 trials required at least one retraction, with 3 trials requiring four. The violin plot in Figure 10b confirms this upward trend, showing an increase in retraction counts as stenosis severity rises. When the experiments were repeated using a more sensitive detection threshold (75 mA, Figure 10c), the number of retractions increased across all stenosis levels. At this lower threshold, the robot initiated retractions even for less severe stenosis. However, the increase in retraction counts was not as pronounced as in the rigid material experiments. Figure 10d presents the corresponding violin plot, illustrating this trend.

**Figure 10.**
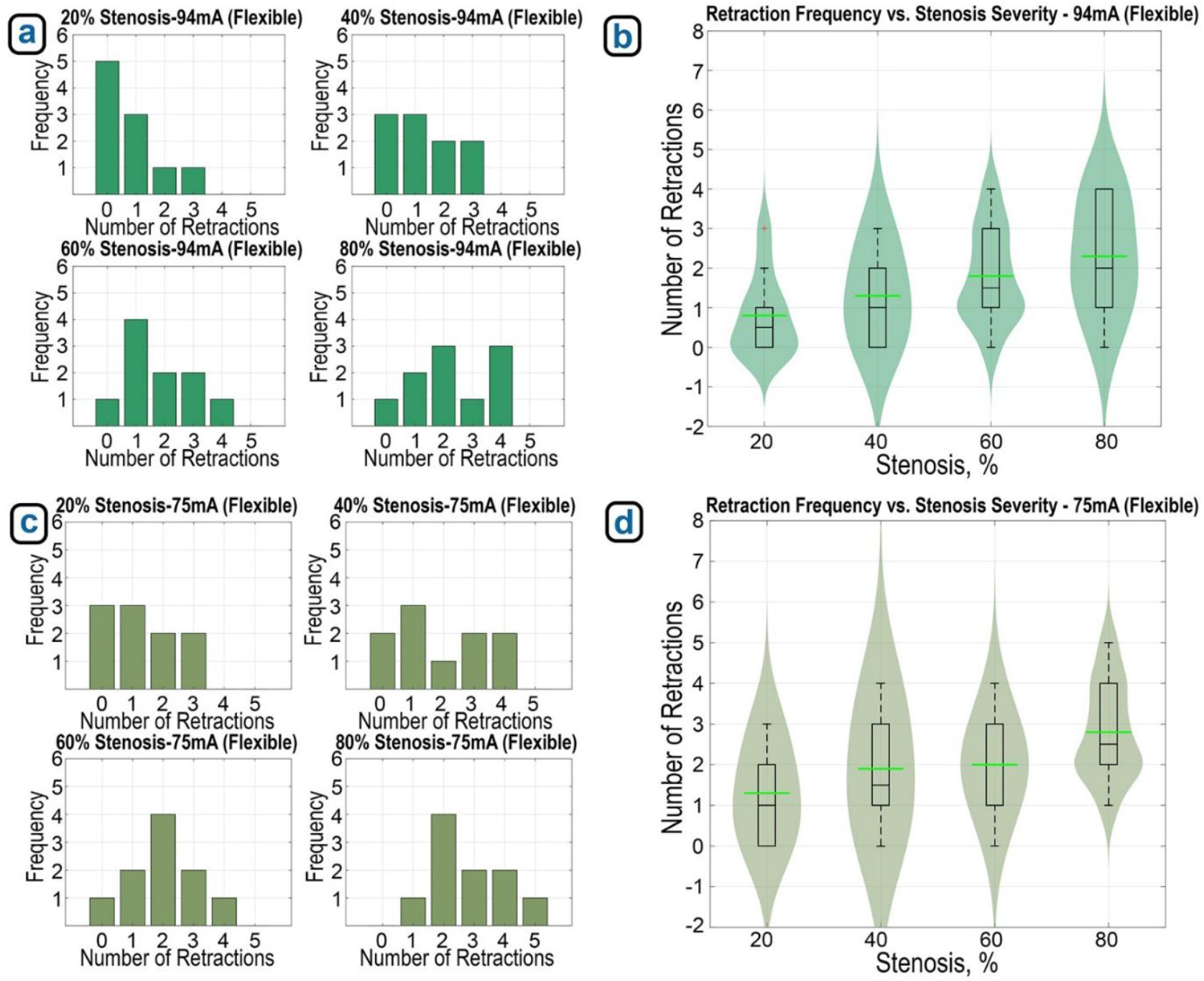
(a) Average number of retractions at each stenosis severity level (flexible, 10 trials each) with 94 mA detection threshold, (b) violin plot showing distribution and density of retractions across stenosis severity under this threshold. (c) Average retractions per stenosis severity (flexible, 10 trials each) at 75 mA threshold, and (d) violin plot showing distribution of retractions across stenosis severity with increased sensitivity.

A three-way ANOVA was conducted to evaluate the effects of stenosis severity (20%, 40%, 60%, 80%), detection sensitivity (94 mA vs. 75 mA), and material stiffness (rigid vs. flexible) on the number of retractions required during guidewire navigation. The analysis revealed that all three main effects were statistically significant. Figure 11 illustrates that retraction counts increased with more severe stenosis (p < 0.001), higher detection sensitivity (p < 0.001), and with stiffer material (p < 0.01). Additionally, a significant interaction between sensitivity and material type was observed (p = 0.031), suggesting that the effect of sensitivity on retraction frequency depends on obstruction stiffness. Furthermore, while increased sensitivity resulted in more retractions for both materials, the increase was more pronounced in rigid obstructions (Figure 11a) compared to the flexible ones (Figure 11b). These results highlight that robot performance is influenced by a combination of physical and control parameters, and that adaptive control strategies may be needed for different material and stenosis contexts.

**Figure 11.**
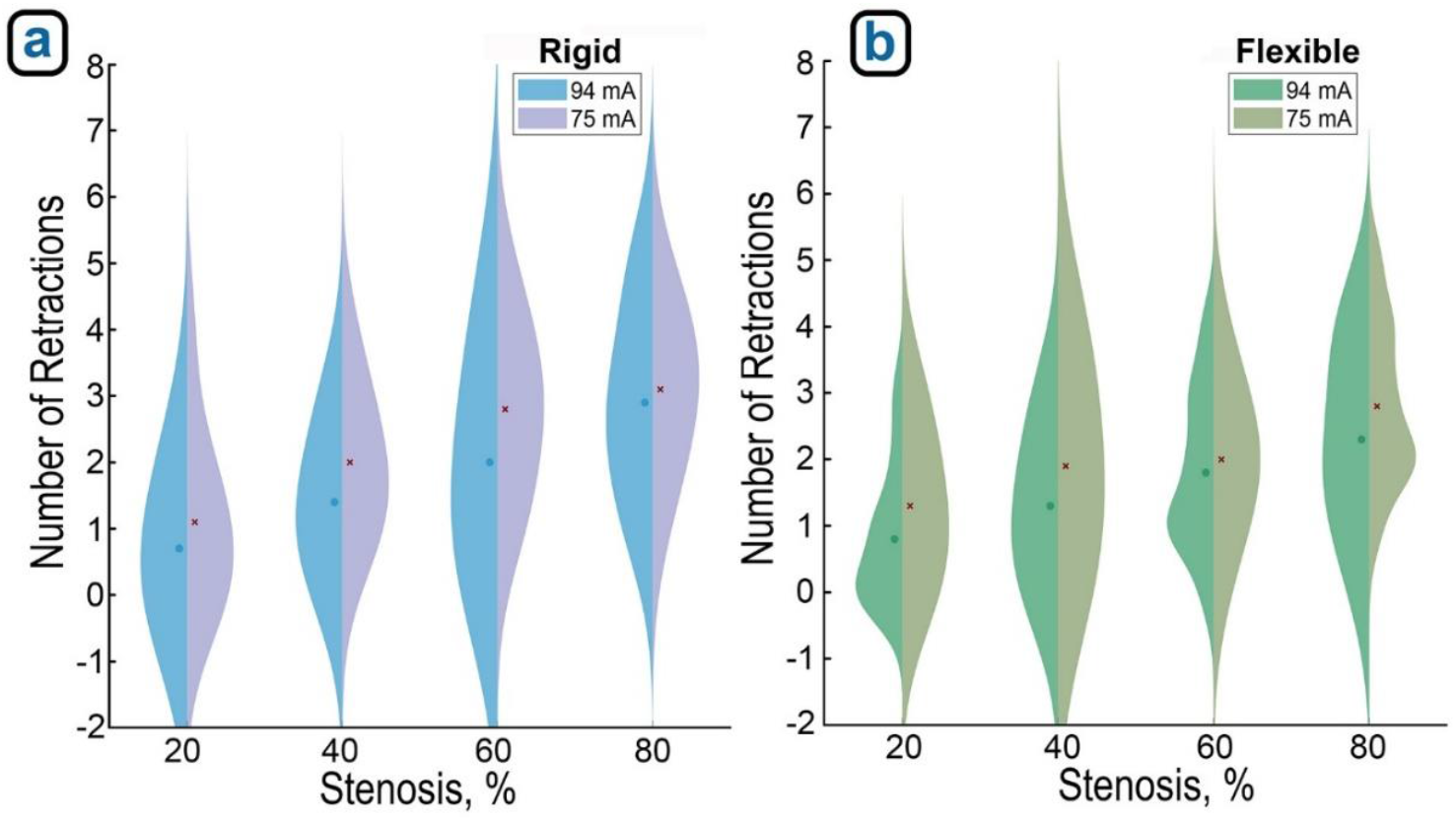
Split violin plots illustrate how retraction counts increase with both stenosis severity and higher detection sensitivity when using (a) rigid and (b) flexible models. Note stronger trends for the rigid obstructions.

### 3.3 Evaluation of Impedance Control and Stiffness Classification

To assess the robot’s impedance control algorithm, we next evaluated its ability to classify obstructions by stiffness, simulating the real-world challenge of distinguishing between soft thrombi and calcified plaques. Three 60% stenosis connectors (Figure 12a) were manufactured with different material properties: stiff (rigid plastic, Formlabs Clear Resin V4.1, Ultimate Tensile Strength 53 MPa, Elongation at Break 9%, 2.5 mm thick), medium (Formlabs Flexible Resin 80A, Ultimate Tensile Strength 8.9 MPa, Elongation at Break 120%, 2.5 mm thick), and soft (Formlabs Flexible Resin 80A, Ultimate Tensile Strength 8.9 MPa, Elongation at Break 120%, 1mm thick).

**Figure 12.**
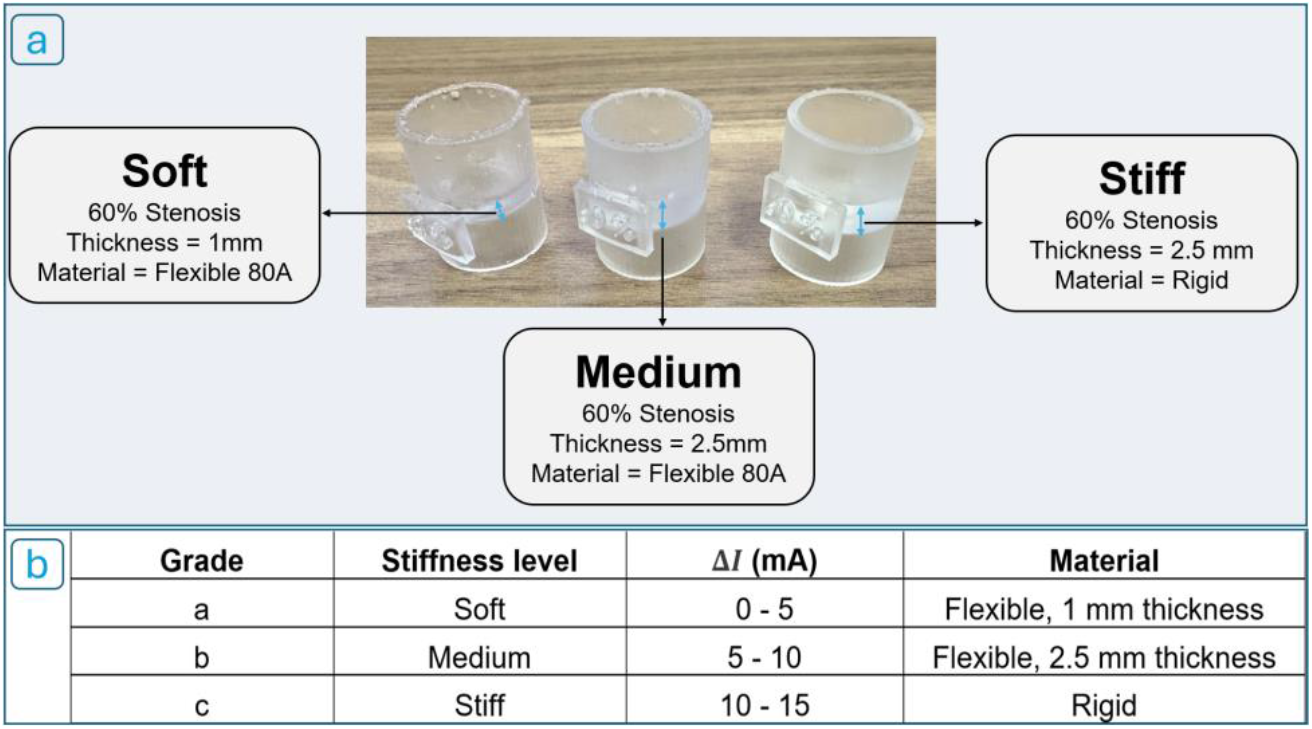
(a) Three 3D-printed stenosis phantoms representing soft, medium, and stiff lesions. (b) Obstruction stiffness classification based on motor current difference (ΔI).

The robot conducted 20 trials per stiffness level, for a total of 60 experiments. Obstruction stiffness was inferred using the difference in motor current before and after a 1 mm advancement, using the formula:

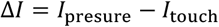

We then classified obstructions into three stiffness grades, based on empirically derived ΔI ranges (Figure 12b).

Figure 13a presents the results of the stiffness classification experiment. The robot correctly identified 19 out of 20 stiff obstructions (95% accuracy), 17 out of 20 soft obstructions (85% accuracy), and 15 out of 20 medium-stiffness obstructions (75% accuracy). Classification performance was highest for stiff lesions and lowest for medium-stiffness cases. These results are further illustrated in the F-score radar chart in Figure 13b which summarizes the precision-recall balance across all three stiffness categories.

**Figure 13.**
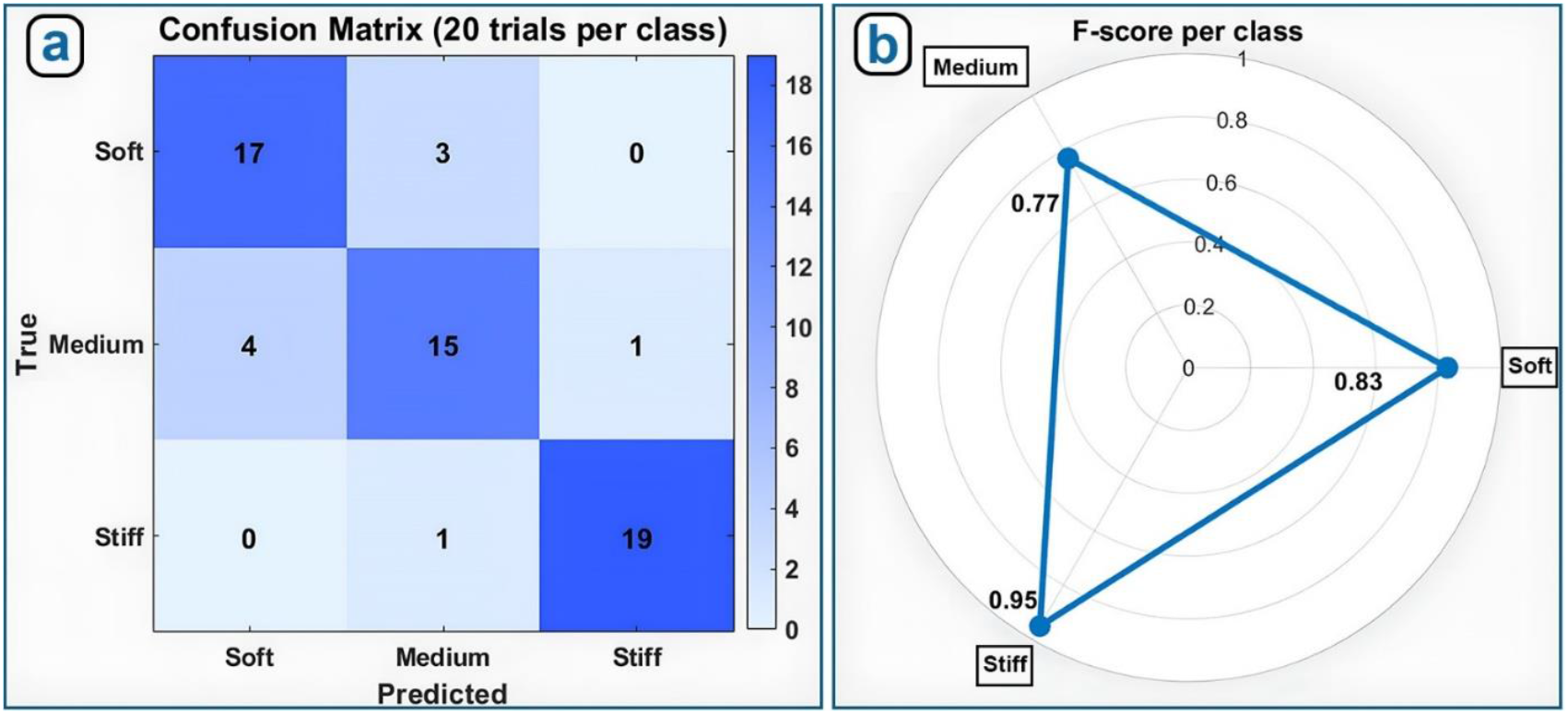
(a) Confusion matrix summarizing correct and incorrect classifications of obstruction stiffness; (b) F-score radar plot depicting classification accuracy for soft, medium, and stiff obstructions.

## 4 DISCUSSION

Robotic automation of endovascular navigation holds immense potential to improve the safety, efficiency, and accessibility of vascular interventions, particularly in time-sensitive or resource-limited settings. In this study, we developed and validated a compact, portable robot ic system that emulates the fine motor skills of an experienced interventionalist - advancing, retracting, and rotating a standard guidewire with millimeter-scale precision. Leveraging internal motor current as a proxy for tactile feedback, the robot adapts its behavior in real time through a layered control strategy combining finite-state logic with impedance-based navigation. The system demonstrated robust performance across a range of stenosis severities in a controlled benchtop model. As stenosis severity increased from 20% to 80%, the number of retractions required to traverse each lesion rose predictably, validating the force-based detection mechanism. Concurrently, the robot successfully classified obstruction stiffness into soft, medium, and stiff categories, achieving F-scores of 0.83, 0.77, and 0.95, respectively. Classification was most accurate for stiff obstructions, which elicited high current responses and minimal displacement, and most challenging for medium-stiffness lesions, which produced ambiguous force profiles. Lowering the detection threshold from 94 mA to 75 mA increased sensitivity but also triggered more retractions, illustrating a trade-off between early detection and procedural efficiency.

In contrast to existing robotic systems, our platform introduces several critical differentiators. Commercial systems such as the Corindus CorPath GRX, Hansen Sensei, and Robocath R-One have established the feasibility of robot-assisted percutaneous interventions, particularly for cardiology and peripheral vascular procedures [33], [34]. However, they were primarily designed for teleoperation, allowing clinicians to manipulate guidewires and catheters from a shielded console to reduce radiation exposure and operator fatigue. Therefore, they still depend on skilled human operators for real-time decision-making and manual navigation. More critically, these platforms are physically large, tethered to fluoroscopic imaging infrastructure, and cost-prohibitive for deployment outside tertiary care centers. Their reliance on hospital-grade equipment and controlled environments makes them unsuitable for pre-hospital, rural, or resource-limited settings where rapid intervention is often most needed. In addition to their infrastructural dependence, most commercial systems lack haptic or force feedback, a crucial feature in traditional manual procedures where interventionalists rely heavily on tactile cues to assess resistance and avoid vessel injury. Without tactile input, robotic systems may advance too forcefully, increasing the risk of dissection or perforation, especially in tortuous or calcified anatomy.

To overcome these challenges, some investigational platforms have proposed to use external force sensors, vision-based feedback, or advanced catheter technologies. Efforts involving machine learning, reinforcement learning, or behavioral cloning have also shown promise in simulations and phantoms [35], [36], [37]. For instance, Song et al. used reinforcement learning and real-time tracking to autonomously control catheter and guidewire navigation through bifurcated vessels[35], while Zhang et al. developed a master-slave robotic catheter system with magnetorheological fluid-based haptic feedback to improve the safety and precision of cardiac ablation procedures in anatomical phantoms under conditions of limited imaging [38]. However, most of these approaches rely heavily on complicated external tracking systems, advanced imaging modalities (e.g., OCT, 3D angiography), or highly specialized catheters, limiting their clinical translatability and increasing cost. Moreover, while some use virtual haptics through force models or torque estimation, true onboard tactile feedback - particularly in a compact, untethered device - has not yet been demonstrated.

Our system represents a significant departure from existing robotic navigation paradigms by combining real-time haptic sensing via motor current, semi-autonomous control logic, and full compatibility with standard endovascular tools in a compact, battery-powered device roughly the size of a smartphone. It operates without external sensors, tracking systems, or imaging infrastructure – reducing cost and complexity while enhancing mechanical robustness. This minimalist design enables reliable use across a wide range of clinical environments, including those far beyond traditional hospital settings. In contrast to handheld vascular access tools such as SAGIV, AI-GUIDE, or VeniBot [39], which are limited to initial vessel entry, our system supports full-length guidewire advancement through tortuous and partially obstructed paths. It can classify lesion stiffness and traverse stenosis of 80%, surpassing the capabilities of most handheld or tethered systems. These features help our platform bridge the gap between bulky, hospital-bound robotic suites and low-autonomy insertion aids, while its compact, self-contained form factor makes it well-suited for deployment in austere environments such as ambulances, rural clinics, battlefield settings, or even space missions - where conventional robotic systems are impractical. In pre-hospital trauma care, it could enable rapid guidewire navigation for procedures like REBOA, where time is critical. Similarly, it may facilitate emergency TEVAR access or serve as a pre-positioning tool for thrombectomy catheters in acute stroke care, helping bridge patients to definitive treatment. Operating without fluoroscopic infrastructure, while still delivering real-time haptic feedback, the robot reduces reliance on expert operators and hospital-grade imaging. Its remote operability also enhances radiation safety by enabling staff to remain outside the fluoroscopy field. Together, these features underscore the system’s versatility and translational potential across a range of time-critical and resource-limited scenarios.

However, despite its promising performance, the current study has several limitations. First, all experiments were conducted in simplified tubing models that approximate vessel geometry but lack the anatomical complexity, compliance heterogeneity, and surface properties of real arteries. The use of a single-lumen tube also precludes evaluation of branch navigation or bifurcation handling. Second, the system was tested in static conditions without fluid flow, which omits important physiological factors such as pulsatile pressure, drag forces, or embolic risk during lesion crossing. Third, the stiffness classification experiments were limited to a fixed stenosis severity and used phantoms with simplified material properties, which may not fully reflect the mechanical behavior of in vivo lesions. Additional testing across a broader range of obstruction geometries and stiffness profiles is needed to validate generalizability. Moreover, while the robot leverages motor current for tactile feedback, it currently lacks true torque sensing, pressure feedback, or imaging integration - capabilities that may be important for navigating tortuous or branching vessels safely. These limitations underscore the need for future validation in anatomically accurate flow models, cadaver studies, and ultimately in vivo systems to assess the system’s performance under more realistic procedural conditions.

## 5 CONCLUSIONS

This study demonstrates that a compact, portable robotic system can safely and reliably guide a standard wire through simulated vascular obstructions. The prototype - approximately the size of a smartphone and weighing 400 grams - was tested in a custom setup that mimics the human vasculature with varying degrees of narrowing. The robot advances, retracts, and rotates the guidewire with millimeter precision, using motor current as a proxy for tactile feedback. This single signal drives a two-layer control system: a finite-state machine that responds in real time to resistance, and an impedance-based controller that probes lesion stiffness and adapts force and speed accordingly. In 80 trials across four stenosis severities (20% to 80% narrowing), the robot successfully navigated all blockages, requiring more retractions as stenosis severity increased - mirroring the behavior of skilled clinicians. In a separate set of 60 trials, the robot accurately classified obstructions as soft, medium, or stiff, achieving F-scores of 0.83, 0.77, and 0.95, respectively. These results suggest several potential clinical benefits: the device is fully portable and battery powered, enabling use in ambulances or rural hospitals; it allows the operator to remain farther from the fluoroscopy source, reducing radiation exposure; and it responds to resistance within milliseconds, potentially lowering the risk of vessel perforation. Future studies will focus on testing the system in anatomically realistic models with blood flow, followed by animal or cadaver validation. Additional sensing modalities such as torque or pressure may further enhance its ability to assess complex plaques. Even in its current form, this lightweight and low-cost platform shows strong potential to expand access to endovascular care and improve the safety and efficiency of vascular procedures.

## Data Availability

All data produced in the present study are available upon reasonable request to the authors

## DECLARATIONS

## Ethics approval and consent to participate Clinical trial number

Not applicable.

## Consent for publication

Not applicable.

## Availability of data and materials

The datasets used and/or analyzed during the current study are available from the corresponding author on reasonable request.

## Authors’ contributions

Conception and design: VM, JM, AK Acquisition of data: VM

Analysis of data: VM, MJ, AK Interpretation of data: VM, AK

Drafting and revising the manuscript: VM, MJ, AK

Approval of the submitted version: VM, JM, MJ, AK

## Acknowledgements

This work was supported in part by the NIH awards HL125736, HL180371, and P20GM152301. The authors would also like to thank the Tissue Analysis Core (TAC) of the NIH Center for Cardiovascular Research in Biomechanics (CRiB) for assistance with experiments.

## REFERENCES

1. Lanza, C., S. Carriero, E. F. M. Buijs, S. Mortellaro, C. Pizzi, L. V. Sciacqua, P. Biondetti, S.A. Angileri, A. A. Ianniello, A. M. Ierardi, and G. Carrafiello. Robotics in Interventional Radiology: Review of Current and Future Applications. Technol. Cancer Res. Treat. 22:15330338231152084, 2023. 10.1177/15330338231152084

2. United States (US) Peripheral Vascular Procedures Count by Segments (Angiography Procedures, Angioplasty Procedures and Others) and Forecast, 2015-2030a <https://www.globaldata.com/store/report/usa-peripheral-vascular-procedures-market-analysis/?utm_source=chatgpt.com>

3. Doll, J. A., A. J. Nelson, L. A. Kaltenbach, D. Wojdyla, S. W. Waldo, S. V. Rao, and T. Y. Wang. Percutaneous Coronary Intervention Operator Profiles and Associations With In-Hospital Mortality. Circ. Cardiovasc. Interv. 15:e010909, 2022. 10.1161/CIRCINTERVENTIONS.121.010909

4. Kamel, H., N. S. Parikh, A. Chatterjee, L. K. Kim, J. L. Saver, L. H. Schwamm, K. S. Zachrison, R. G. Nogueira, O. Adeoye, I. Díaz, A. M. Ryan, A. Pandya, and B. B. Navi. Access to Mechanical Thrombectomy for Ischemic Stroke in the United States. Stroke 52:2554–2561, 2021. 10.1161/STROKEAHA.120.033485

5. Eckardt, L., F. Doldi, O. Anwar, N. Gessler, K. Scherschel, A.-K. Kahle, A. S. Von Falkenhausen, R. Thaler, J. Wolfes, A. Metzner, C. Meyer, S. Willems, J. Köbe, P. S. Lange, G. Frommeyer, K.-H. Kuck, S. Kääb, G. Steinbeck, and M. F. Sinner. Major in-hospital complications after catheter ablation of cardiac arrhythmias: individual case analysis of 43 031 procedures. europace 26:euad361, 2023. 10.1093/europace/euad361

6. Caicedo, Y., L. M. Gallego, H. Jc. Clavijo, N. Padilla-Londoño, C.-N. Gallego, I. Caicedo-Holguín, M. Guzmán-Rodríguez, J.J. Meléndez-Lugo, A. F. García, A. E. Salcedo, M. W. Parra, F. Rodríguez-Holguín, and C.A. Ordoñez. Resuscitative endovascular balloon occlusion of the aorta in civilian pre-hospital care: a systematic review of the literature. eur. J. Med. Res. 27:202, 2022. 10.1186/s40001-022-00836-3

7. Harper, C., S. A. Collier, and T. L. Slesinger. Traumatic Aortic Injuries. In: StatPearls. Treasure Island (FL): StatPearls Publishing, 2025.at <http://www.ncbi.nlm.nih.gov/books/NBK555980/>

8. Brede, J. R., T. Lafrenz, P. Klepstad, E. A. Skjærseth, T. Nordseth, E. Søvik, and A.J. Krüger. Feasibility of Pre-Hospital Resuscitative Endovascular Balloon Occlusion of the Aorta in Non-Traumatic Out-of-Hospital Cardiac Arrest. J. Am. Heart Assoc. 8:e014394, 2019. 10.1161/JAHA.119.014394

9. Witberg, G., V. Tzalamouras, H. Adams, T. Patterson, R. Roberts-Thomson, J. Byrne, R. Dworakowski, P. MacCarthy, S. Redwood, and B. Prendergast. Routine Ultrasound or Fluoroscopy Use and Risk of Vascular/Bleeding Complications After Transfemoral TAVR. JACC Cardiovasc. Interv. 13:1460–1468, 2020. 10.1016/j.jcin.2020.03.047

10. Eo, S. J., D. S. Ryu, C. E. Yun, Y. Park, D.-S. Won, J. W. Kim, S. H. Kim, J.-H. Park, and D.-H. Kim. One-hand guidewire introducer kit for ultrasound-guided central venous catheterization: a proof-of-concept study. Sci. Rep. 14:29222, 2024. 10.1038/s41598-024-79784-3

11. Duan, W., T. Akinyemi, W. Du, J. Ma, X. Chen, F. Wang, O. Omisore, J. Luo, H. Wang, and L. Wang. Technical and Clinical Progress on Robot-Assisted Endovascular Interventions: A Review. Micromachines 14:197, 2023. 10.3390/mi14010197

12. Yang, D., J. Song, and Y. Hu. Guidewire feeding method based on deep reinforcement learning for vascular intervention robot., 2022. 10.1109/ICMA54519.2022.9856351

13. Scarponi, V., M. Duprez, F. Nageotte, and S. Cotin. A Zero-Shot Reinforcement Learning Strategy for Autonomous Guidewire Navigation., 2024. 10.48550/ARXIV.2403.02777

14. Zhang, B., M. Bui, C. Wang, F. Bourier, H. Schunkert, and N. Navab. Real-time guidewire tracking and segmentation in intraoperative x-ray., 2022. 10.1117/12.2611097

15. Okamura, A. M. Haptic feedback in robot-assisted minimally invasive surgery: Curr. Opin. Urol. 19:102–107, 2009. 10.1097/MOU.0b013e32831a478c

16. Pescio, M., D. Kundrat, and G. Dagnino. Endovascular robotics: technical advances and future directions. Minim. Invasive Ther. Allied Technol. 1–14, 2025. 10.1080/13645706.2025.2454237

17. Payne, C. J., H. Rafii-Tari, and G.-Z. Yang. A force feedback system for endovascular catheterisation., 2012. 10.1109/IROS.2012.6386149

18. Yang, X., H. Wang, L. Sun, and H. Yu. Operation and force analysis of the guide wire in a minimally invasive vascular interventional surgery robot system. Chin. J. Mech. eng. 28:249–257, 2015. 10.3901/CJME.2014.1229.181

19. Lesaunier, A., J. Khlaut, C. Dancette, L. Tselikas, B. Bonnet, and T. Boeken. Artificial intelligence in interventional radiology: Current concepts and future trends. Diagn. Interv. Imaging 106:5–10, 2025. 10.1016/j.diii.2024.08.004

20. Lu, Q., Z. Sun, J. Zhang, J. Zhang, J. Zheng, and F. Qian. A Novel Remote-Controlled Vascular Interventional Robotic System Based on Hollow Ultrasonic Motor. Micromachines 13:410, 2022. 10.3390/mi13030410

21. Da, L., D. Zhang, and T. Wang. Overview of the vascular interventional robot. Int. J. Med. Robot. 4:289–294, 2008. 10.1002/rcs.212

22. Ravigopal, S. R., T. A. Brumfiel, and J. P. Desai. Automated Motion Control of the COAST Robotic Guidewire under Fluoroscopic Guidance., 2021. 10.1109/ISMR48346.2021.9661508

23. Baltrūnas, T., V. Labunskas, V. Dambrauskas, E. Kalvaitis, and V. Bajoras. Smart and Sensing Endovascular Robot Sentante. J. Vasc. Surg. 75:16S, 2022. 10.1016/j.jvs.2022.01.059

24. Schegg, P., J. Dequidt, E. Coevoet, E. Leurent, R. Sabatier, P. Preux, and C. Duriez. Automated Planning for Robotic Guidewire Navigation in the Coronary Arteries., 2022. 10.1109/RoboSoft54090.2022.9762096

25. Baek, H., B. Cheon, J. M. You, and D.-S. Kwon. Design and analysis of feeder mechanism for buckling prevention in robotic catheterization. J. Comput. Des. eng. 9:1467–1481, 2022. 10.1093/jcde/qwac065

26. Lee, C. J.-Y., J. L. Pallisgaard, J. B. Olesen, N. Carlson, M. Lamberts, G. H. Gislason, C. Torp-Pedersen, A. Brandes, S. E. Husted, S. P. Johnsen, and M. L. Hansen. Antithrombotic Therapy and First Myocardial Infarction in Patients With Atrial Fibrillation. J. Am. Coll. Cardiol. 69:2901–2909, 2017. 10.1016/j.jacc.2017.04.033

27. Siracuse, J. J., and A. Farber. Is Open Vascular Surgery or Endovascular Surgery the Better Option for Lower Extremity Arterial Occlusive Disease? Adv. Surg. 51:207–217, 2017. 10.1016/j.yasu.2017.03.016

28. Bdiwi, M., I. Al Naser, J. Halim, S. Bauer, P. Eichler, and S. Ihlenfeldt. Towards safety4.0: A novel approach for flexible human-robot-interaction based on safety-related dynamic finite-state machine with multilayer operation modes. Front. Robot. AI 9:1002226, 2022. 10.3389/frobt.2022.1002226

29. Alkkiomäki, O., V. Kyrki, H. Kälviäinen, Y. Liu, and H. Handroos. Complementing visual tracking of moving targets by fusion of tactile sensing. Robot. Auton. Syst. 57:1129–1139, 2009. 10.1016/j.robot.2009.07.001

30. Song, P., Y. Yu, and X. Zhang. A Tutorial Survey and Comparison of Impedance Control on Robotic Manipulation. Robotica 37:801–836, 2019. 10.1017/S0263574718001339

31. Mohammadi, V., R. Shahbad, M. Hosseini, M. H. Gholampour, S. Shiry Ghidary, F. Najafi, and A. Behboodi. Development of a Two-Finger Haptic Robotic Hand with Novel Stiffness Detection and Impedance Control. Sensors 24:2585, 2024. 10.3390/s24082585

32. Hogan, N. Impedance Control: An Approach to Manipulation., 1984. 10.23919/ACC.1984.4788393

33. Mahmud, E., F. Schmid, P. Kalmar, H. Deutschmann, F. Hafner, P. Rief, and M. Brodmann. Feasibility and Safety of Robotic Peripheral Vascular Interventions. JACC Cardiovasc. Interv. 9:2058–2064, 2016. 10.1016/j.jcin.2016.07.002

34. Cha, H.-J., H.-S. Yoon, K. Y. Jung, B.-J. Yi, S. Lee, and J. Y. Won. A robotic system for percutaneous coronary intervention equipped with a steerable catheter and force feedback function., 2016. 10.1109/IROS.2016.7759194

35. Song, H.-S., B.-J. Yi, J. Y. Won, and J. Woo. Learning-based catheter and guidewire-driven autonomous vascular intervention robotic system for reduced repulsive force. J. Comput. Des. eng. 9:1549–1564, 2022. 10.1093/jcde/qwac074

36. Garg, S., M. Goharimanesh, S. Sajjadi, and F. Janabi-Sharifi. Autonomous control of soft robots using safe reinforcement learning and covariance matrix adaptation. eng. Appl. Artif. Intell. 153:110791, 2025. 10.1016/j.engappai.2025.110791

37. Fagogenis, G., M. Mencattelli, Z. Machaidze, B. Rosa, K. Price, F. Wu, V. Weixler, M. Saeed, J. E. Mayer, and P. E. Dupont. Autonomous robotic intracardiac catheter navigation using haptic vision. Sci. Robot. 4:eaaw1977, 2019. 10.1126/scirobotics.aaw1977

38. Zhang, L., J. Zuo, K. Wang, T. Jiang, S. Gu, L. Xu, and Y. Zhang. An advanced robotic system incorporating haptic feedback for precision cardiac ablation procedures. Sci. Rep. 15:6839, 2025. 10.1038/s41598-025-91342-z

39. Brattain, L., T. Pierce, L. Gjesteby, M. Johnson, N. DeLosa, J. Werblin, J. Gupta, A. Ozturk, X. Wang, Q. Li, B. Telfer, and A. Samir. AI-Enabled, Ultrasound-Guided Handheld Robotic Device for Femoral Vascular Access. Biosensors 11:522, 2021. 10.3390/bios11120522

